# Relative expression of pro-inflammatory molecules in COVID-19 patients manifested disease severities

**DOI:** 10.1101/2021.04.01.21254770

**Authors:** Shireen Nigar, SM Tanjil Shah, Md. Ali Ahasan Setu, Sourav Dutta Dip, Habiba Ibnat, M Touhidul Islam, Selina Akter, Iqbal Kabir Jahid, M Anwar Hossain

## Abstract

Aggressive immune response, due to over-expressed pro-inflammatory molecules, had been characterized in COVID-19 patients. Some of those mediators have a dual and opposite role on immune-systems to play behind differential disease severities. We investigated the expression of some cytokines and chemokines in COVID-19 patients in Bangladesh. We diagnosed the patients by detecting SARS-CoV-2 RNA in nasal swab samples by the real-time RT-PCR method. Thirty adult patients were preselected based on their disease severities and grouped into mild, moderate, and severe cases. Nine healthy volunteers participated in this study as control. Relative expression of nine cytokines/chemokine in total leukocytes was semi-quantified in SYBRgreen-based qRT-PCR. We performed statistical tests on transformed log data using SPSS 24.0. At the onset of symptoms (day-1), ACE2 (P < 0.05) and IL-6 (P > 0.05) were up-regulated in all COVID-19 groups, although expression levels did not significantly correlate with disease severities. However, expression of IL-6, MCP-1, MIP-1α, TNF-α, RANTES, and ACE2, on day-14, were positively correlated with disease severities. Relative viral load at day-1 showed no significant correlation with cytokine expression but had a significant positive correlation with RANTES and ACE2 expression on day-14 (P < 0.05). Male patients had a higher level of IL-6 than female patients on day-1 (P < 0.05). All COVID-19 patients showed up-regulated cytokines and chemokines on the day-14 compared to day-1 except TNF-α. Female patients had higher expression of ACE2 and IL-12 on day-14. Up-regulated cytokines/chemokines at the convalescent stage, especially IL-6, may target anti-cytokine therapy in post-COVID-19 patients’ management.

## Introduction

A novel coronavirus SARS-CoV-2 was identified in Wuhan, China, in December 2019 and has lead to over 127 million cases and 2.79 million death worldwide as of March 31, 2021 ^[1]^. However, surprisingly in Bangladesh, these days recorded 0.61 million confirmed cases and only 9046 deaths ^[1]^. COVID-19 related deaths are significantly lower in countries with lower quality of life ^[2]^. Although most SARS-CoV-2 patients demonstrate mild or moderate symptoms, it could also lead to excessive, ineffective and exaggerated immune responses called cytokine release syndrome (CRS). The clinical response shows acute respiratory distress syndrome (ARDS), multiple organ failure, and ultimately death ^[3]^. Several authors reported the role of cytokines and chemokines as a double-edged sword. Higher levels of chemokines (such as MCP-1, MIP-1α, MIP-1β, RANTES, IP-10, and CCL3) and cytokines (such as IL-6, IL-8, IL-1b, TNF-α, IFNγ, and GCSF, GMCSF) have reported for COVID-19 patients ^[3-6]^. Several authors have been reviewed the importance and function of these chemokines and cytokines ^[7-9]^ and demonstrated the role of IL-6, TNF-α, IL-10, and IL-8 for disease severity.

The SARS-CoV-2 virus uses novel Metallo-carboxyl peptidase angiotensin receptor 2 (ACE2) to enter its human host cell ^[10, 11]^. In spite of acting as the viral receptor, ACE2 protects against lung injury by degrading vasoconstrictive and proapoptotic protein ^[12]^. ACE2 expressed higher in severe patients than mild and moderate ^[13]^. The authors also found upregulation of several interferons, cytokines, and immune-related genes for severe patients compared with mild and moderate patients. Biological sex variation influenced the severity of COVID-19 cases ^[14]^ as reported in various studies; men are more likely to get severe forms of COVID-19 ^[15-17]^.

Investigations will identify therapeutic strategies with anti-cytokine targeting overactive pro-inflammatory cytokines (TNF-α and IL-6). But the correct time for administrating anti-cytokine therapies needs to be recognized for personalized treatment of COVID-19 patients. Furthermore, the questions remained unanswered about the reasons for reduced severity and mortality in developing countries like Bangladesh, except for dissimilarity in cytokine/chemokine expression with recently reported data. We aimed to explore the relationships between disease severity and relative cytokine expression levels, the effect of biological sex in clinical presentation or viral load at the nasopharynx swap of COVID-19 patients.

## Materials & Methods

### Ethical permission and Sample collection

The Ethical Review Committee of the Jashore University of Science and Technology approved this study (ERC no: ERC/FBST/JUST/2020-49). A total of 30 symptomatic COVID-19 patients were preselected, and patients were categorized into mild cases (n=10), moderate cases (n=10), and severe cases (n=10) of COVID-19 according to the World Health Organization Guidelines for Diagnosis and Treatment of COVID-19 infection ^[18]^. We grouped patients with fever and slight upper respiratory tract symptoms as mild cases; patients with shortness of breathing, constant pain, or pressure in the chest as moderate cases; and patients with respiratory failure requiring intensive care units (ICU) as severe COVID-19 cases. We informed individual participants (COVID-19 patients) or their family members about the study protocol and took verbal consent. The demographic information of all participants enlisted in Table 1. Ten healthy individuals (gave their consent and participated in this study) were primarily selected, who were free from recent respiratory diseases, acute or chronic infectious diseases, and had no co-morbidities. They maintained home-quarantine from the beginning of the COVID-19 outbreak in Bangladesh and tested negative for recent SARS-CoV-2 infection by using All Check COVID-19 IgG/IgM antibody assay (CALTH Inc., Republic of Korea). We exclude a sample from one volunteer in the final study due to the appearance of antibodies against SARS-CoV-2. All participants were BCG vaccinated within one month of birth (identified by the skin scar/mark on the left upper arm and their verbal confirmation). We have interviewed patients or their family members to collect demographic data and clinical history from all qualified participants. From hereafter, we refer to the healthy control as HC, mild COVID-19 cases as MIC, moderate COVID-19 cases as MOC, and severe COVID-19 cases as SC.

**Table 1.** Demographic and clinical characteristics of studied participants.

| Characteristics | HC | MIC | MOC | SC |
| --- | --- | --- | --- | --- |
|  | (n=9) | (n=10) | (n=10) | (n=10) |
| Men | 5 (55.55%) | 6 (60%) | 8 (80%) | 6 (60%) |
| Women | 4 (44.45%) | 4 (40%) | 2 (20%) | 4 (40%) |
| Age [mean(±SD)] years | 23.89(±2.435<br>) | 37.30(±4.814) | 40.20(±4.69) | 58.40(±3.3<br>61) |
| Fever (temperature $>37^{\circ}\text{C}$) | 0 | 8 (80%) | 10 (100%) | 9 (90%) |
| Cough | 0 | 4 (40%) | 8 (80%) | 8 (80%) |
| Headache | 0 | 5 (50%) | 6 (60%) | 6 (60%) |
| Malaise | 0 | 2 (20%) | 10 (100%) | 7 (70%) |
| Myalgia | 0 | 2 (20%) | 5 (50%) | 3 (30%) |
| Loss of taste or odor | 0 | 0 | 6 (60%) | 4 (40%) |
| Shortness of breathing | 0 | 2 (20%) | 7 (70%) | 10 (100%) |
| Constant pain or pressure in chest | 0 | 3 (30%) | 5 (50%) | 5 (50%) |
| SPO<sub>2</sub> at rest $\leq 94\%$ | 0 | 0 | 0 | 6 (60%) |
| Respiratory failure requiring<br>intensive care unit treatment | 0 | 0 | 0 | 10 (100%) |
| Diabetes | 0 | 0 | 0 | 4 (40%) |
| Hypertension | 0 | 0 | 1 (10%) | 6 (60%) |

Abbreviations: HC; Healthy control, MIC; mild COVID-19 cases, MOC; moderate COVID-19 cases, SC; severe COVID-19 cases.

### Quantification of SARS-CoV-2 viral load

We performed detection of SARS-CoV-2 at the Genome Centre, Jashore University of Science and Technology, Jashore, Bangladesh. We extracted viral RNA from a 10 µL clinical specimen (nasal swab and throat swab) within 12 h of specimen collection using QuickExtract™ RNA extraction kit (Lucigen, Wisconsin, USA) following the manufacturer’s instruction. Then, 10 µL of each viral RNA extract was amplified by one-step real-time quantitative reverse-transcriptase polymerase chain reaction (qRT-PCR) using Novel Coronavirus Nucleic Acid Diagnostic Kit (Sansure Biotech Inc., China). The kit detected *n*- and *orf1b* gene of SARS-CoV-2 and human *RNase P* gene as an internal control. In this study, samples with Ct values for both n- and orf1b genes of ≤35.0 considered positive, that of >35.0 retested, and of >40.0 considered negative. We used equation 2^-ΔCt^ to estimate viral load, where ΔCt = Ct _viral N gene_ – Ct_RNaseP_ ^[19]^. We have translated all the data as Log_10_(1+2^-ΔCt^).

### Estimating cytokine and chemokine expression levels by qRT-PCR

We collected approximately 4 to 5 ml of peripheral blood samples, within 24-hours (day 1) of laboratory confirmation of SARS-CoV-2 in nasopharyngeal swabs, from thirty COVID-19 symptomatic and nine symptomatic patients. After 14 days, we collected blood samples from twenty patients with MIC (n=10) and MOC (n=10) symptoms from their residence while maintaining home quarantine. Three healthy volunteers (out of nine) tested SARS-CoV-2 positive during repeated sampling and let off from the study. So we had six samples from healthy volunteers on day-14. We could not collect blood samples from participants of the severe COVID-19 cases as they were either decreased after 14 days of initial sample collection or admitted into the intensive care unit (ICU) from whom we could not be allowed to collect the blood.

We extracted total mRNA from human leukocytes using 1 mL of whole blood from all study participants using the SV total RNA isolation system (Promega, USA), as per the manufacturer’s instruction. According to the manufacturer’s instruction, we prepared 20.0 µL cDNA from 8.0 µL of total mRNAs using the GoScript™Reverse transcription system (Promega, USA). We diluted 20 µL cDNA into a final volume of 100.0 µL. We performed SYBRgreen intercalation qRT-PCR to detect the expression level of two pro-inflammatory cytokines (TNF-α and IL-6), two antiviral cytokines (IFN-γ and IL-12), four chemokines (MIP-1α, RANTES, IP-10, and MCP-1), and ACE2. We used the primer sequences to detect these cytokines and chemokines from an earlier study ^[20]^. The qRT-PCR was performed in QuantStudio™ 3 (Applied Biosystems, USA), using the following protocol: denaturation at 95 °C for 2 min followed by 40 cycles of 95 °C for 10 s, annealing for 30 s, and extension at 72 °C for 30 s. We analyzed the melting curve to determine the assay’s specificity (60-95 °C, 0.05°C/s). Annealing temperature varied for different primer pairs (such as ACE2: 53 °C; IL-6: 45 °C; TNF-α: 60 °C; IP-10: 45 °C; IFN-γ: 48 °C; MIP-1α: 58 °C; RANTES: 48 °C; MCP-1: 52 °C; IL-12: 50.5 °C; and β-actin: 60 °C). We performed each experiment in triplicate and Ct values with <10% variance considered for analyses only. In this essay, the human β–actin gene worked as the internal control. We accessed mRNA expression by relative quantification and calculated fold expression change by the2^-ddCt^ method ^[21]^.

### Statistical analysis

The distribution of the fold expression change of mRNAs was skewed, so we log-transformed these data for further analyses. Results presented as median (interquartile range). Non-parametric tests such as the Kruskal-Walis test and Mann-Whitney U-Test were used to compare relative cytokine expression levels among the different groups where applicable. Spearman’s rank correlation coefficient (r) analyzes the correlation between relative cytokine expression levels in total lymphocytes and the close viral load. Spearman rank correlation analyses the association between disease severity and cytokine expression levels. We performed statistical tests based on transformed log data using SPSS 24.0 for Windows (SPSS, Inc., Chicago, IL, USA) and GraphPad Prism 8.0 (GraphPad Software, USA) to construct all the figures logarithmic scales. If the two-tailed P values were <0.05, the test results were considered significant.

## Result

### Demographic and clinical characteristics of studied participants

The demographic characteristics and clinical aspects of thirty COVID-19 patients and nine healthy volunteers listed in Table 1. Mean(±SD) age of MIC, MOC, and SC groups were estimated as 37.30(±4.814), 40.20(±4.69), and 58.40(±3.361) years, respectively. HC groups’ mean age was 23.89(±2.435) years. All MOC participants (n=10) and SC (n=10) suffered from shortness of breathing, but only two MIC reported such difficulties. All SC had respiratory failure and thus required ICU support. We recorded no death for mild and moderate groups but one death reported during the study period in the SC group.

### Relative expression levels of cytokine and chemokine in studied participants

Fig 1 represented the comparison between different cytokine and chemokine expression levels on day-1 and day-14 among the study groups. Statistical analyses showed no significant difference (P > 0.05) among the four groups for any cytokine or chemokine on day-1, except for ACE2. The ACE2 expressed higher (P < 0.05) in the MIC group than HC group. The relative expression level of IL-6 was numerically higher in all COVID-19 groups than in the healthy control group, although this difference was non-significant (P > 0.05).

**Figure 1:**
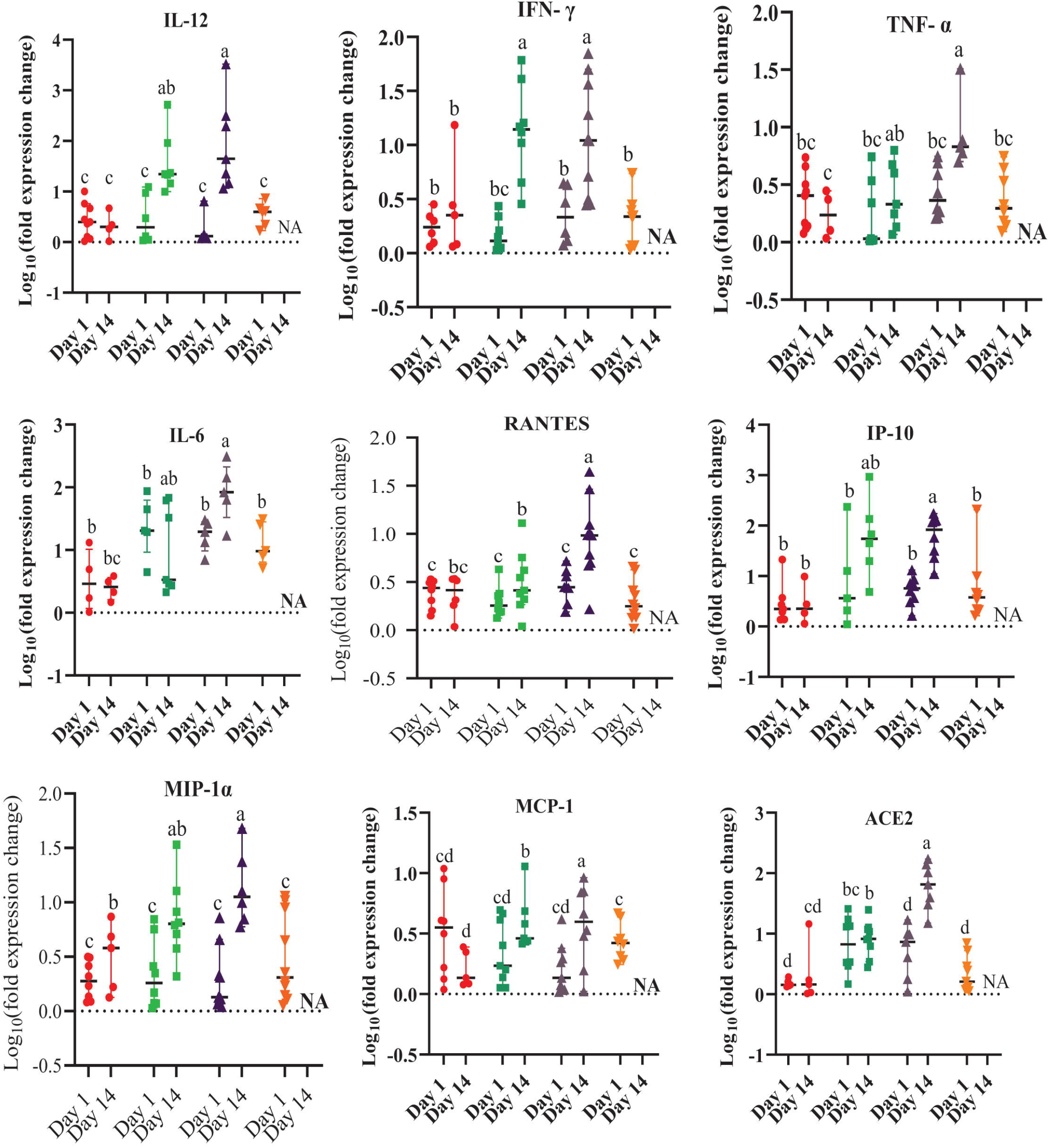
Relative expression levels of cytokine and chemokine in healthy control and COVID-19 infected symptomatic patients (mild COVID-19 cases, moderate COVID-19 cases, and severe COVID-19 cases). ● = Healthy Control; ■ = Mild COVID-19 cases; ▲ = Moderate COVID-19 cases; and ▼ = Severe COVID-19 cases. Each RT-PCR was done in triplicate and data those vary <10% were taken for analyses. Because of this, number of samples varies for different cytokine and chemokine. Data are expressed as median with interquartile range (IQR). Separate analyses were performed for day 1 and day 14. For any cytokine, different letters in different column indicates significant differences (P < 0.05). Abbreviations: NA, not available (severe COVID-19 cases data on day 14); IL, interleukin; IFN, interferon; TNF, tumor necrosis factor; RANTES, Regulated upon activation, normal T cell expressed and secreted; IP-10, interferon-inducible protein of 10 kDa; MIP, macrophage inflammatory protein; MCP, monocyte chemotactic protein; and ACE, angiotensin converting enzyme.

To obtain changes of different cytokine and chemokine expression levels on day-14. Compared with the HC, the MIC group had increased (P < 0.05) expressions of IL-12 and IFNγ and non-significant higher expression of IP-10 and MCP-1 at day-14. On the other hand, the MOC had statistically higher (P < 0.05) expressions of ACE2, IL-12, MCP-1, TNF-α, MIP-1α, IL-6, RANTES, IP-10, and IFN-γ on day-14 compared to the HC. Compared with the MIC group, the MOC had numerically higher IL-6 levels and significantly higher expression (P < 0.05) of RANTES and ACE2.

### Correlation between disease severity and cytokine and chemokine expression level

Spearman rank correlation analyses were performed to determine any correlations between disease severity and expression levels of cytokine and chemokine in COVID-19 patients, separately for both day-1 and day-14. Correlation coefficients (r) for all cytokine and chemokine (both day-1 and day-14) are shown in Fig 2A. Statistical analyses revealed that the relative expression levels of IL-6 (r= 0.609), TNF-α (r= 0.741), MIP-1α (r= 0.607), MCP-1 (r= 0.486), RANTES (r= 0.623) and ACE2 (r= 0.845) on day-14 were positively correlated (P < 0.05) with disease severity (Fig 2B). However, no significant correlation between disease severity with any cytokine and chemokine was observed on day-1. But expression levels of IL-12 (r= 0.102), TNF-α (r= 0.110), IFN-γ (r= 0.166), IL-6 (r= 0.295), IP-10 (r= 0.210), and MIP-1α (r= 0.128) were weakly positively correlated with severity on day-1.

**Figure 2:**
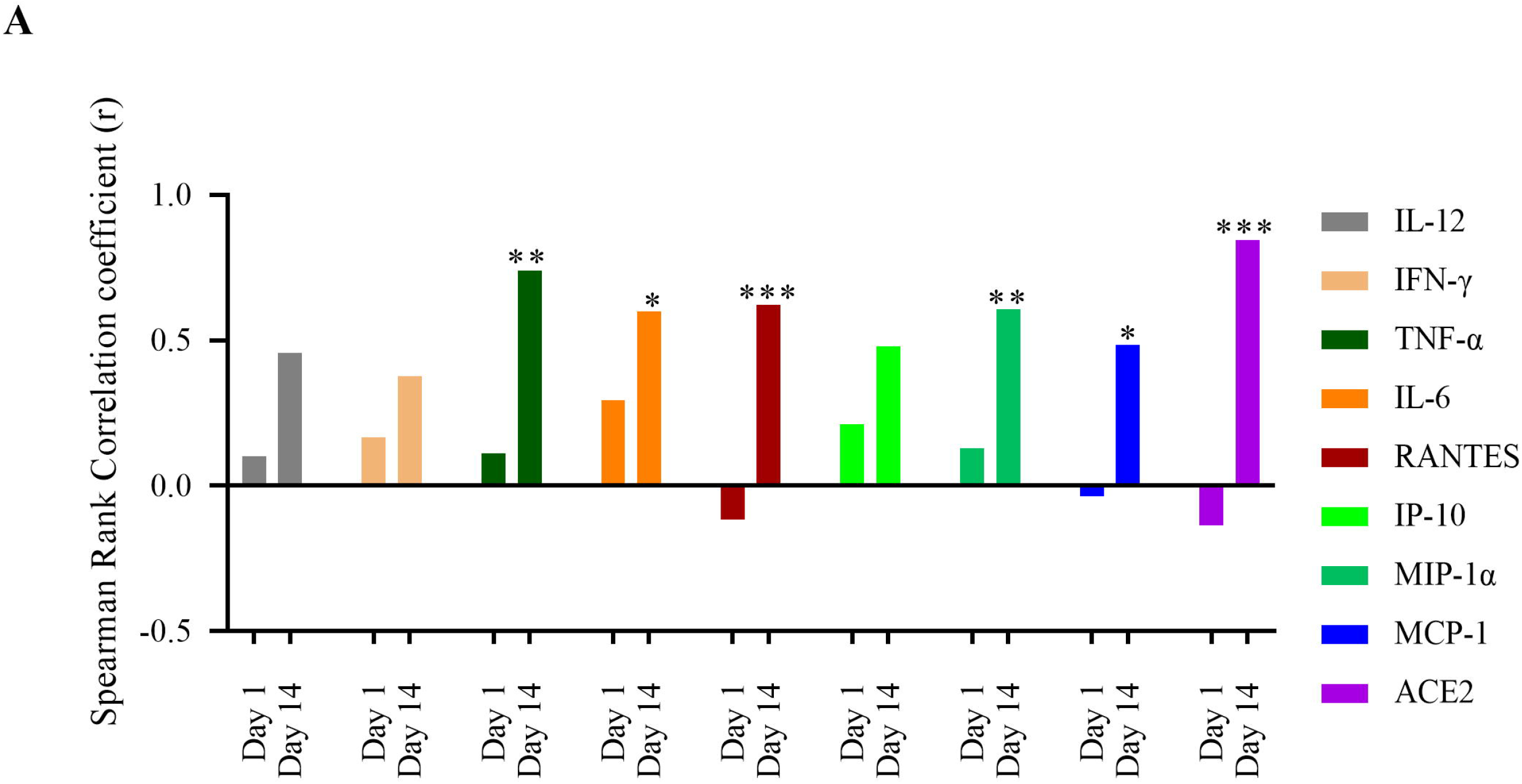
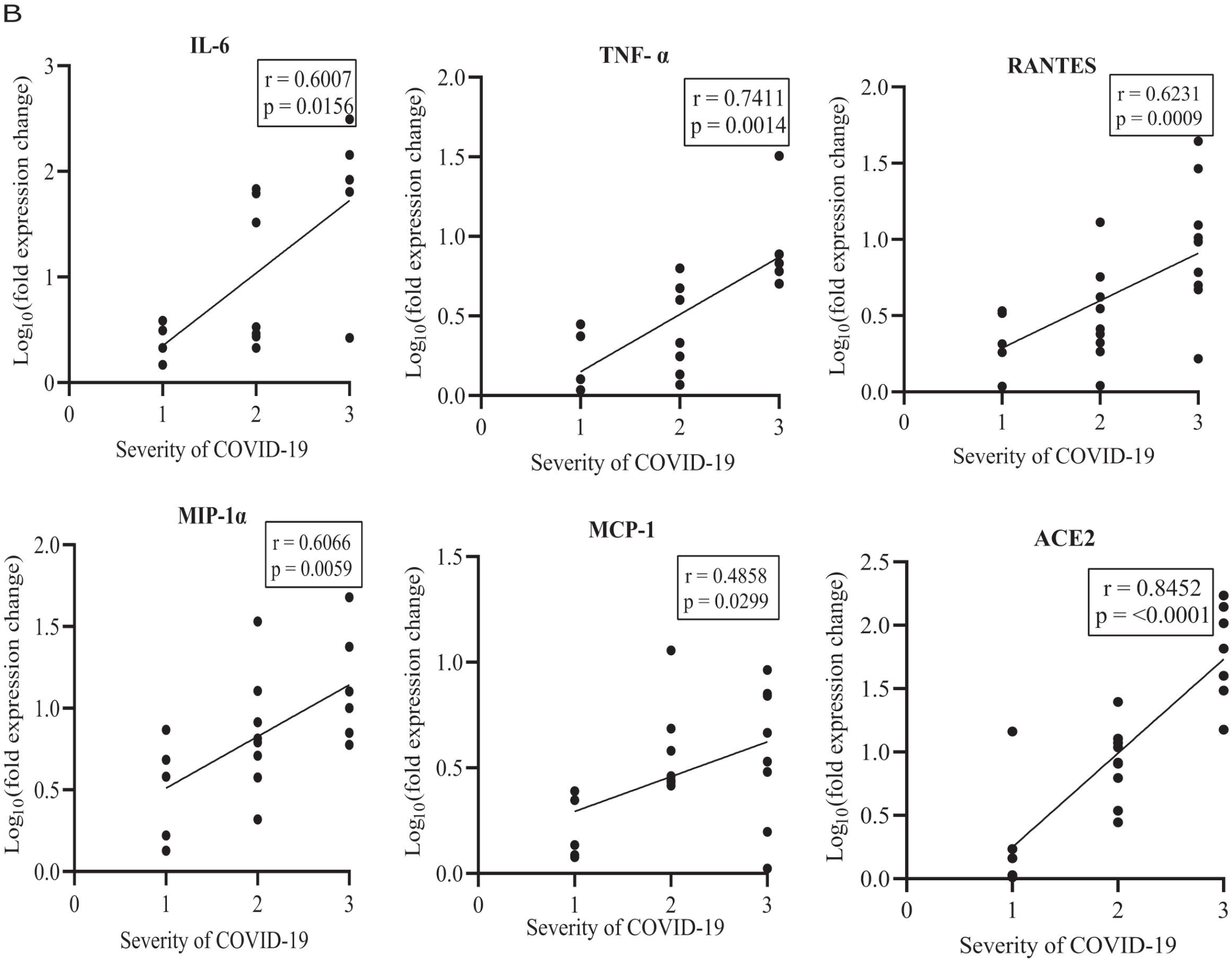
Correlation between disease severity and relative expression levels of cytokine and chemokines. (A).Spearman rank correlation analyses between disease severity and cytokine expression levels were done, and correlation coefficient (r) values of IL-12, IFN-γ, TNF-α, IL-6, RANTES, IP-10, MIP-1α, MCP-1, and ACE2 on day 1 and day 14 are plotted. * P < 0.05, **P < 0.01, ***P < 0.001. (B). Correlation between disease severity and relative expression levels of IL-6, TNF-α, RANTES, MIP-1α, MCP-1, and ACE2 after 14 days are plotted. Here in the X axis, 1= healthy volunteer, 2= mild COVID-19 cases, and 3= moderate COVID-19 cases.Abbreviations: IL, interleukin; IFN, interferon; TNF, tumor necrosis factor; RANTES, Regulated upon activation, normal T cell expressed and secreted; IP-10, interferon-inducible protein of 10 kDa; MIP, macrophage inflammatory protein; MCP, monocyte chemotactic protein; and ACE, angiotensin converting enzyme.

### Correlation between viral load and cytokine and chemokine expression level

Spearman rank correlation analyses were also done to determine any correlations between relative viral load and expression levels of cytokine and chemokine in COVID-19 patients. Correlation coefficients (r) for all cytokine and chemokine (both day 1 and day 14) are shown in Fig 3A. Expression levels of only TNF-α (r = 0.4469) had non-significant positive correlation (P > 0.05) with relative viral load at the onset of disease. Whereas IL-6 (r= −0.4182), RANTES (r = −0.2376), MIP-1α (r = −0.2078), and MCP-1 (r = −0.3111) had non-significant weak negative correlation with relative viral load on day 1. However after 14 days, expression levels of RANTES (r = 0.4860) and ACE2 (r = 0.6471) had significant positive correlation (P < 0.05) with the relative viral load (Fig 3B). Moreover, relative expression levels were non-significantly positively correlated with viral load for IL-12 (r = 0.3516), TNF-α (r = 0.4615), IL-6 (r = 0.4895), and MIP-1α (r = 0.3538).

**Figure 3:**
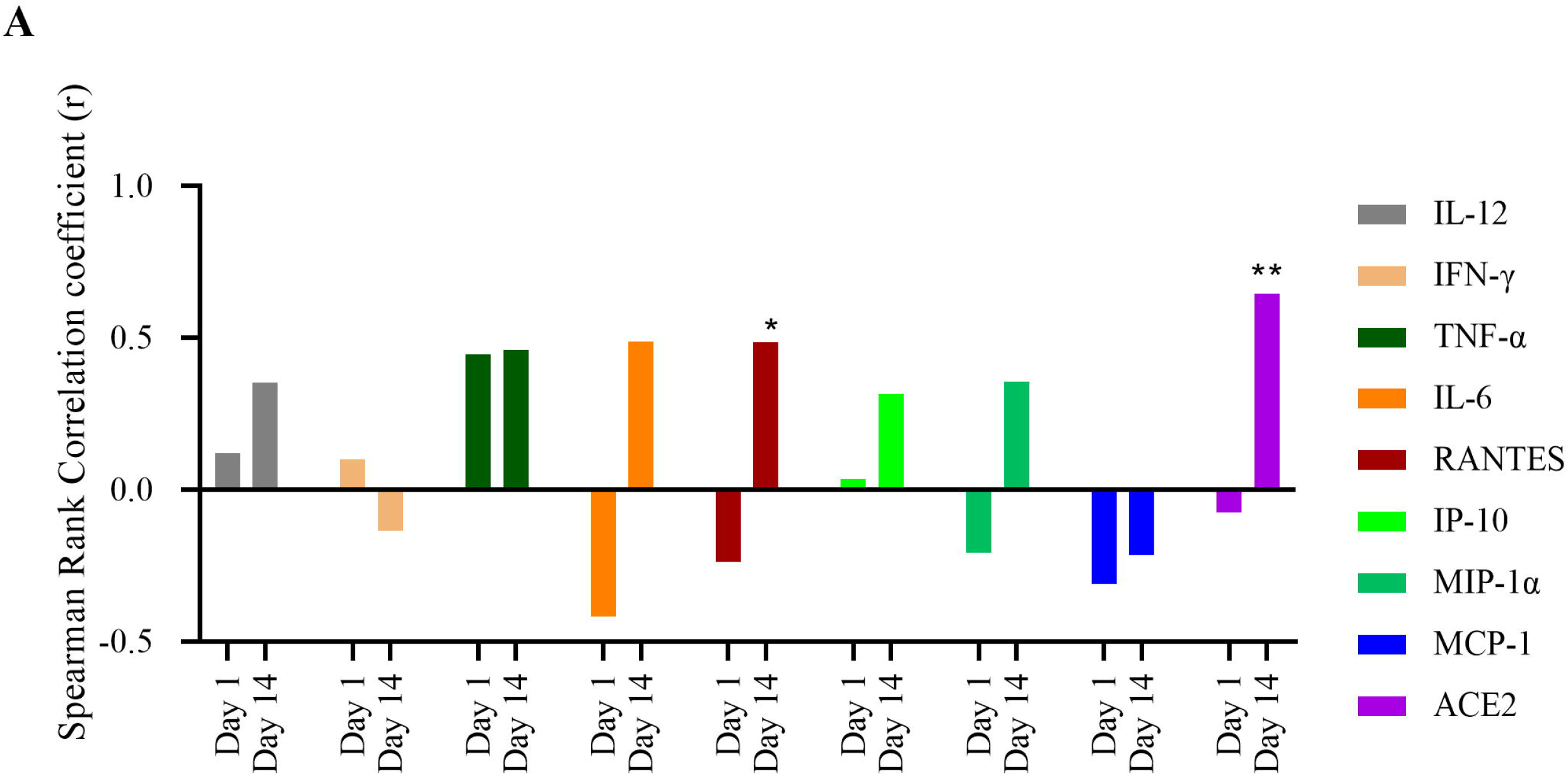
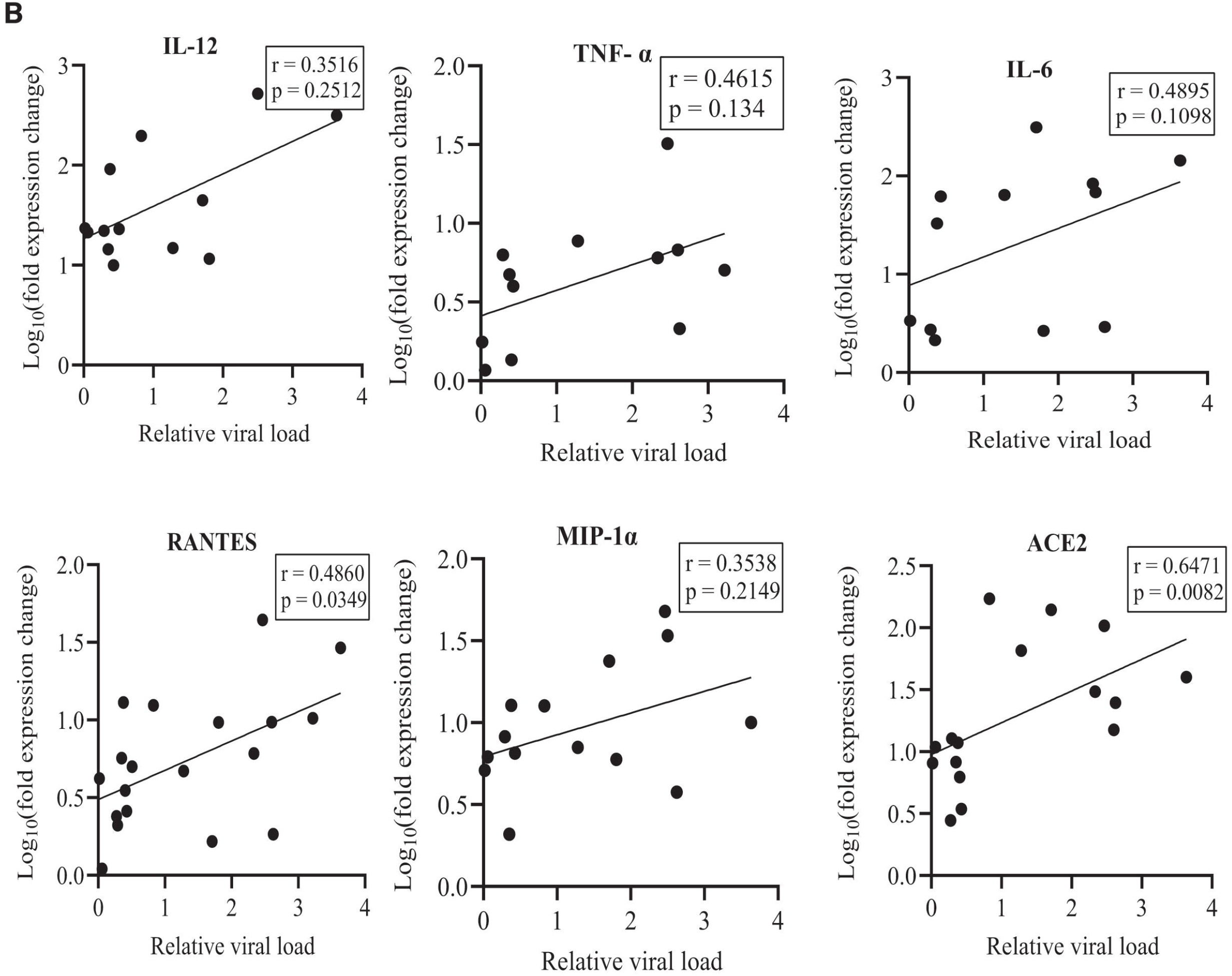
Correlation between relative viral load and relative expression levels of cytokine and chemokines in COVID-19 patients. (A). Spearman rank correlation analyses between relative viral load and cytokine expression levels were done, and correlation coefficient (r) values of IL-12, IFN-γ, TNF-α, IL-6, RANTES, IP-10, MIP-1α, MCP-1, and ACE2 on day 1 and day 14 are plotted. * P < 0.05, **P < 0.01. (B). Correlation between relative viral load and expression levels of IL-12, TNF-α, IL-6, RANTES, MIP-1α, and ACE2 after 14 days are plotted. Abbreviations: IL, interleukin; IFN, interferon; TNF, tumor necrosis factor; RANTES, Regulated upon activation, normal T cell expressed and secreted; IP-10, interferon-inducible protein of 10 kDa; MIP, macrophage inflammatory protein; MCP, monocyte chemotactic protein; and ACE, angiotensin converting enzyme.

### Effect of gender on relative cytokine expression levels in COVID-19 patients

To explore any effect of sex on cytokine and chemokine expression levels, we analyzed cytokine’s relative expression levels of males and females on both day-1 and day-14 (Fig 4). Irrespective of sex, all cytokine and chemokine were up-regulated on day-14 than day-1, except for IL-6 and TNF-α. For male, IL-6 expression level was similar on both sampling times. TNF-α showed no significant difference in any comparison. The relative expression level of ACE2 and IL-12 were also identical in both gender on day-1. But both ACE2 and IL-12 were up-regulated (P < 0.05) in females on day-14. Additionally, we did not observe a significant difference in the relative expression level of IFN-γ, RANTES, and IP-10 in males and females on both days. On the other hand, MCP-1 and MIP-1α were up-regulated (P < 0.05) in females on day-1. But did not found any sex discrimination for expression of MCP-1 and MIP-1α on day-14. The relative expression level of IL-6 was higher (P < 0.05) in males on day-1 but did not vary significantly (P > 0.05) with genders on day-14.

**Figure 4:**
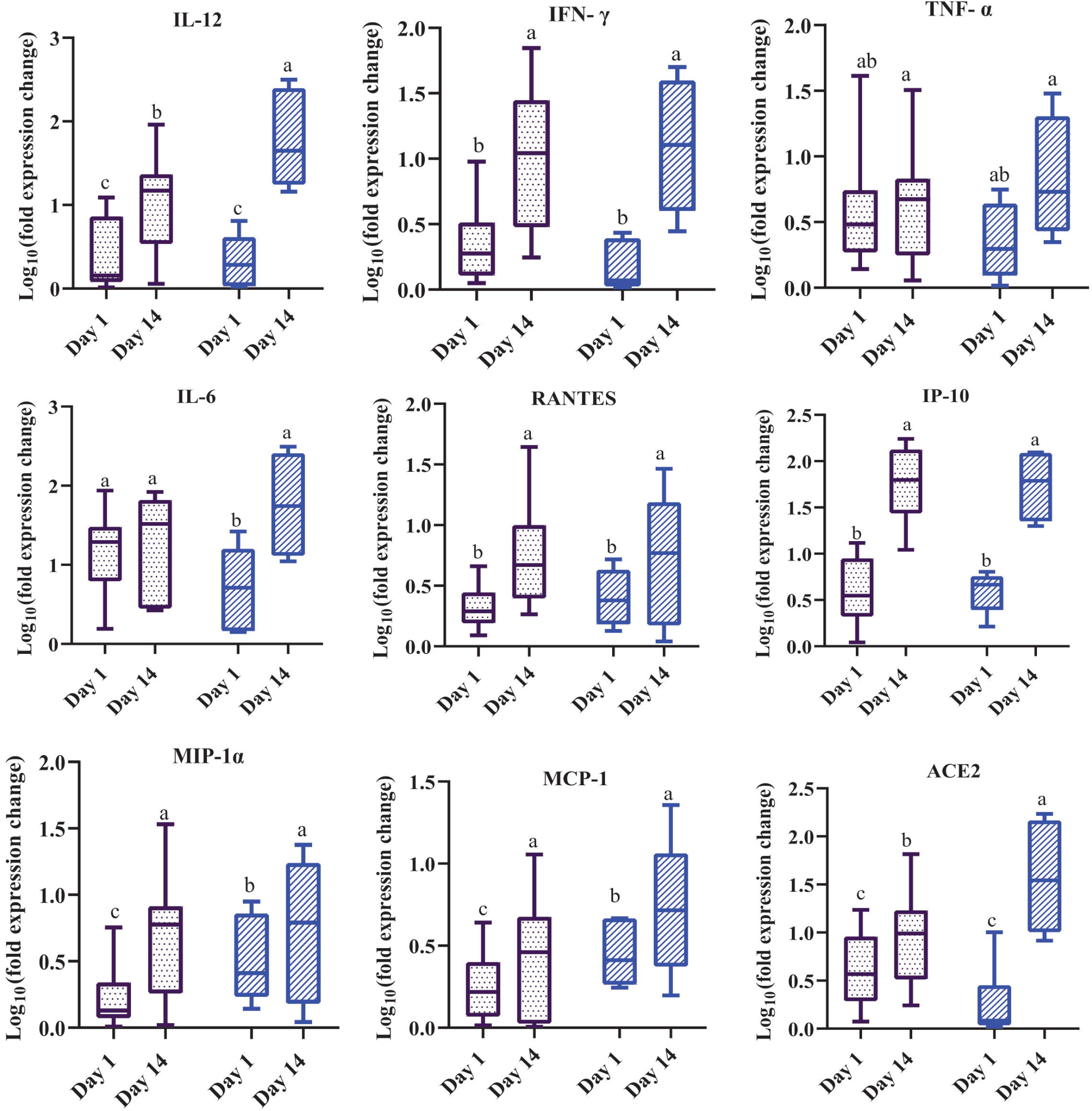
Relative expression levels of cytokine and chemokine in male and female COVID-19 patients. Here, ▭= male and ▭= female COVID-19 infected patients. Each RT-PCR was done in triplicate and data those vary <10% were taken for analyses. Because of this number of samples varies for different cytokine and chemokine. Data are expressed as median with interquartile range (IQR). Separate analyses were performed for day 1 and day 14. For any cytokine, different letters in different column indicates significant differences (P < 0.05). Abbreviations: IL, interleukin; IFN, interferon; TNF, tumor necrosis factor; RANTES, Regulated upon activation, normal T cell expressed and secreted; IP-10, interferon-inducible protein of 10 kDa; MIP, macrophage inflammatory protein; MCP, monocyte chemotactic protein; and ACE, angiotensin converting enzyme.

## Discussion

To our knowledge, this is the first study on cytokine/chemokine expression level in COVID-19 patients in Bangladesh. We observed up-regulation of ACE2 (P < 0.05) and IL-6 (P > 0.05) in MIC cases compared with HC on day-1 (Fig 1). Many natural, common ways such as taking Vitamin C, Metformin, Vitamin B3, and Vitamin D can up-regulate ACE2 receptor ^[22]^. Here in Bangladesh, doctors instructed COVID-19 patients to take Vitamin C and Vitamin B3 during their illnesses, increasing ACE2 expression. Although the virus uses ACE2 to enter into its host cell, up-regulation of the cellular ACE2 receptor would likely be anti-inflammatory and might have contributed to reduced mortality in COVID-19 patients. Pro-inflammatory cytokines such as IL-6, TNF-α, while up-regulated, trigger cytokine storms and ultimately lead to tissue damage and organ failure ^[23]^. Although we observed a slightly increased expression of IL-6, we didn’t observe TNF-α up-regulation at the onset of the disease (Fig 1). Like our study, normal TNF-α levels were reported in COVID-19 patients by Wan et al. ^[24]^. However, elevated TNF-α levels in COVID-19 patients’ sera were also reported recently ^[4, 25]^. Recently some studies reported IL-6 as severity predictors of COVID-19 ^[26, 27]^. In a meta-analysis including 1426 patients, complicated COVID-19 patients had higher IL-6 expression levels (>3 times) compared with other uncomplicated patients ^[28]^. In our study, COVID-19 patients had no higher expression of either IFN-γ or IL-12 than the HC group, which is indicative of the absence of elevated antiviral immune response at the onset of disease symptoms. Surprisingly, none of the cytokines and chemokines of ICU patients from the SC group shows significant differences compared with others. Zhang et al ^[29]^ found that ICU patients in China had higher levels of IL-2R, IL-6, IL8, IL10, and TNF-α compared with reference ranges of the normal man. In other studies from Finland, ICU patients had higher levels of IL-6, C-reactive protein, and procalcitonin compared with non-ICU patients ^[30]^. Keddie et al ^[31]^ also noted similar findings. The authors demonstrated that CRP, IL-6, IL-10, and LDH significantly correlated with ARDS and respiratory support. In another study, Lev et al ^[32]^ also noted that IP-10 is associated with ICU patients and act as potential biomarkers. The combination of IL-6*IL-10 serum levels has shown a predictor of ICU patients ^[33]^. The less expression of most cytokines and chemokines might be probable reasons for less mortality rate (1.5%) of severe COVID-19 patients in Bangladesh.

Moreover, in our study, the relative expression of cytokine and chemokines differs between 2 weeks. After 14 days, significant up-regulation of both antiviral cytokines (IFN-γ and IL-12) occurs in MIC and MOC. Although the antiviral immune response was not present in COVID-19 patients on the onset of disease, the antiviral immune response accelerated in those patients after 14 days. This up-regulation indicates enhanced host immunity to eliminate viral pathogens. Similar IL-12 up-regulation reported in both SARS-CoV ^[34]^ and SARS-CoV-2 infected patients ^[4, 35]^. IL-12 has distinctive characteristics and has a vital role in positive and negative feedback ^[36]^. On day 14, pro-inflammatory cytokine IL-6 was significantly higher in MOC but not in MIC cases. Whereas another pro-inflammatory cytokine, TNFα, was significantly up-regulated in all COVID-19 patients than HC. These findings suggest that pro-inflammatory cytokines had direct effects on the severity and complexity of COVID-19 disease. The anti-inflammatory cytokines levels got up-regulated compared to pro-inflammatory cytokines after 14 days. So, it might result in rapid recovery and reduced mortality in COVID-19 patients. On day-14, RANTES and ACE2 also up-regulated with the severity of COVID-19 disease. RANTES, a well-established chemo-attractant, links innate and adaptive immune response. RANTES decreased significantly in SARS-CoV-2 infected persons in a recent study, where MIP-1α level was higher in ICU patients ^[4]^.

We observed that the relative expression of some cytokines and chemokines varied among COVID-19 cases. Based on the samples’ data on day 1, we could not establish any significant correlation between disease severity and relative cytokine expressions. But after 14 days, estimated expression levels of IL-6, TNF-α, MIP-1α, MCP-1, RANTES, and ACE2 correlated with and differed significantly with COVID-19 severity groups (Fig 2). Several reviews highlighted the importance of cytokines and chemokines and demonstrated IL-1, IL-6, IL-8, IL-10, IL-2R, IL-18, MCP-1, and TNF-α for disease severity ^[8, 37, 38]^. The cytokines and chemokines regulate the CD4 + T, CD8 +T, monocytes, neutrophils, and macrophages. However, we observed differences in the expressions between studies. Recent work by Qin et al ^[25]^ is also in accord with our observation, where COVID-19 severity was associated with increased levels of IL-6, IP-10, MCP-1, and MIP-1α. Although Costela-Ruiz et al ^[7]^ reported elevated TNF-α and its association with severity in COVID-19 patients, we didn’t observe such differences. In another study, even the samples from nasopharyngeal swabs revealed the higher expression of IFN-γ and lowered expression of TGF-β1 and RANTES in symptomatic patients compared with negative patients ^[39]^. But our studies demonstrated the higher expression of RANTES for MIC and MOC compared with HC. At the onset of disease, the relative viral load had no significant correlation with cytokines and chemokines’ relative expression (Fig 3A). However, initial viral load positively correlated with expression levels of IL-12, TNF-α, IL-6, RANTES, MIP-1α, and ACE-2, after 14 days. A higher level of IL-6 and RANTES decreased CD8+ T cell and virus ^[40]^. The results from our study agreed with that study.

Although male tends to get more severe forms of COVID-19 diseases, we observed significant gender variation only for IL-6, MIP-1α, and MCP-1 on day 1 (Fig 4). Our observations of male patients who had up-regulated IL-6 than female patients at the beginning of the disease agreed with China’s similar findings. The study reported a higher level of serum IL-6 male compared to female. Increased expression of IL-6 on day-1 might result in severe COVID-19 diseases in males. Interestingly both males and females had similar expression levels of pro-inflammatory cytokines (TNF-α, IL-6) after 14 days. On day-14, female patients had significant up-regulation of ACE2 and IL-12. In response to vaccinations, women induced a robust immune response ^[41]^. In a previous study, estrogens lead to cytokine up-regulation in mice treated with coronavirus. Estrogens had a protective role by suppressing the immune response’s escalation phase ^[42]^. As women are expressing more ACE2, therefore, less severity and death rate has been attributed in Bangladesh. A recent study showed that, compared with male patients, severe female patients had a greater IgG production level in weeks of the COVID-19 disease onset ^[6]^. The protective role of IgG in female COVID-19 patients might be associated with the up-regulation of IL-12.

There were a few limitations along with the small sample size in our study. We could not collect repeated blood samples after 14 days from severe patients admitted to ICU. We also estimated the relative expression levels of cytokines and chemokines instead of serum cytokine levels. However, live vaccines such as the bacillus Calmette-Guerin (BCG) are known to induce trained immunity (enhanced innate immune response to subsequent infections). COVID-19 cases reported being lower in countries with universal BCG vaccination programs (such as Bangladesh, Nepal, Bhutan, Japan) compared to those without the programs (such as the USA, Spain, Canada, Italy) ^[43]^. We hypothesized that BCG vaccination might induce an immune response and reduce SARS-CoV-2 viremia and the severity of COVID-19 infections ^[44]^, and rapid recovery of SARS-CoV-2 infection in the country.

## Conclusion

This study describes cytokine and chemokine expression among COVID-19 patients with different disease severity in a developing country, Bangladesh. We found that IL-6 could be targeted for anti-cytokine therapy here in Bangladesh. Although we could not prove any direct effect of BCG vaccinations on reduced severity of COVID-19, our data provide cytokine expression levels in BCG vaccinated COVID-19 patients. Knowledge of the underlying mechanisms of differential expressions and associations of these cytokines with disease severity could help target the choice of therapies.

## Data Availability

The data will be available on request to author

## Acknowledgments

This work was supported by a research grant (UGC/JUST/2020-21/20) from the University Grant Commission, Bangladesh, through Jashore University of Science and Technology (JUST), Jashore. We thank all volunteers and COVID-19 patients who actively participated in this study. We also acknowledge the efforts of all doctors, nurses, health-care workers in fighting SARS-CoV-2.

## Authors’ contribution statement

Shireen Nigar conceived the idea; SM Tanjil Shah, Ali Ahsan Setu, Sourav Datta Dip, Habiba Ibnat and Selina Akter performed the experiment; M Touhidul Islam collected and performed initial processing of samples; SM Tanjil Shah and Iqbal Kabir Jahid performed statistical analysis; Shireen Nigar and SM Tanjil Shah wrote the draft manuscript; Selina Akter, Iqbal Kabir Jahid and M Anwar Hossain finalize the manuscript; Shireen Nigar, Iqbal Kabir Jahid and M Anwar Hossain supervised the whole work and submitted the manuscript.

